# SGLT2 Inhibitors in Stage 5 Chronic Kidney Disease: A Systematic Review and Meta-analysis of Cardiovascular and Kidney Outcomes

**DOI:** 10.64898/2026.11.12.26367903

**Authors:** Hafiz M. Ahmed, Ubaid ur Rehman, Muhammad Owais

## Abstract

Evidence supporting SGLT2 inhibitor use in stage 5 chronic kidney disease remains limited because landmark trials have largely excluded this population. In this systematic review and meta-analysis, SGLT2 inhibitor use was associated with fewer heart failure hospitalizations and myocardial infarctions, while benefits for mortality and dialysis initiation remained uncertain. Dedicated randomized trials are needed to clarify their role in this population.

## 1. INTRODUCTION

Sodium–glucose cotransporter-2 (SGLT2) inhibitors reduce kidney disease progression and heart failure events in patients with an estimated glomerular filtration rate (eGFR) ≥20 mL/min/1.73 m^2^ ^1^. Current guidelines recommend initiating treatment at this threshold and continuing therapy as eGFR declines, but evidence supporting initiation in stage 5 chronic kidney disease (CKD; eGFR <15 mL/min/1.73 m^2^) remains limited ^2^. Previous meta-analyses have primarily included patients with eGFR ≥20 mL/min/1.73 m^2^. More recent observational studies have examined SGLT2 inhibitor use in stage 5 CKD, with mixed findings that have not been quantitatively synthesized. We therefore systematically reviewed the available evidence to assess clinical outcomes and examine the methodological limitations of these studies.

## 2. METHODS

This systematic review followed PRISMA 2020 guidelines and was registered in PROSPERO (CRD420261364520) ^3^. We searched PubMed, Scopus, Cochrane CENTRAL, and Google Scholar from inception through April 10, 2026 **(Table S1)**. Eligible studies compared SGLT2 inhibitor use with nonuse or other glucose-lowering therapies in adults with stage 5 CKD and reported mortality, cardiovascular, renal, or safety outcomes.

Meta-analysis was limited to studies reporting outcomes specifically in stage 5 CKD (eGFR <15 mL/min/1.73 m^2^ or an equivalent diagnostic code) without overlapping populations. Two Taiwanese cohorts used the National Health Insurance Research Database over overlapping periods. To avoid double-counting participants, we included only the larger target trial emulation in the meta-analysis and retained the other study for qualitative synthesis ^4,5^. Another study included patients with CKD stages 3B–5 but reported subgroup outcomes only for eGFR <20 mL/min/1.73 m^2^ ^6^. Because stage 5-specific estimates were unavailable, this study was included in the qualitative synthesis but not pooled.

Two reviewers (H.M.A. and U.R.) independently screened records, extracted study characteristics and outcomes, and assessed risk of bias using ROBINS-I ^7^. Disagreements were resolved through discussion or consultation with a third reviewer (M.O.). We pooled four outcomes: all-cause mortality, dialysis initiation, acute myocardial infarction, and heart failure hospitalization. Log hazard ratios (HRs) were combined using inverse-variance random-effects models, with between-study variance estimated by restricted maximum likelihood (REML) and Wald-type 95% confidence intervals (CIs). Sensitivity analyses used Hartung–Knapp-adjusted confidence intervals and restricted the analysis to the two treatment-initiation cohorts (Yen and Chen), excluding Anuforo because the timing of treatment initiation was unclear. Certainty of evidence was assessed using GRADE. Safety outcomes were summarized narratively because definitions and reporting varied across studies. Publication bias was not assessed because too few studies were available. All analyses were performed in R version 4.5.2.

## 3. RESULTS

Of 1,466 records screened, five observational cohorts met the inclusion criteria, three of which were included in the meta-analysis **(Table 1, Figure S1)** ^4–6,8,9^. The pooled studies included 57,156 participants, of whom 28,559 received SGLT2 inhibitors. Follow-up ranged from a median of 0.9 years to a maximum of 5 years. Four studies compared SGLT2 inhibitor use with nonuse, while one used dipeptidyl peptidase-4 (DPP-4) inhibitors as the comparator. Four studies excluded patients receiving dialysis at baseline, whereas Chen (2025) included 11.5% receiving dialysis in each treatment arm.

**Table 1.**
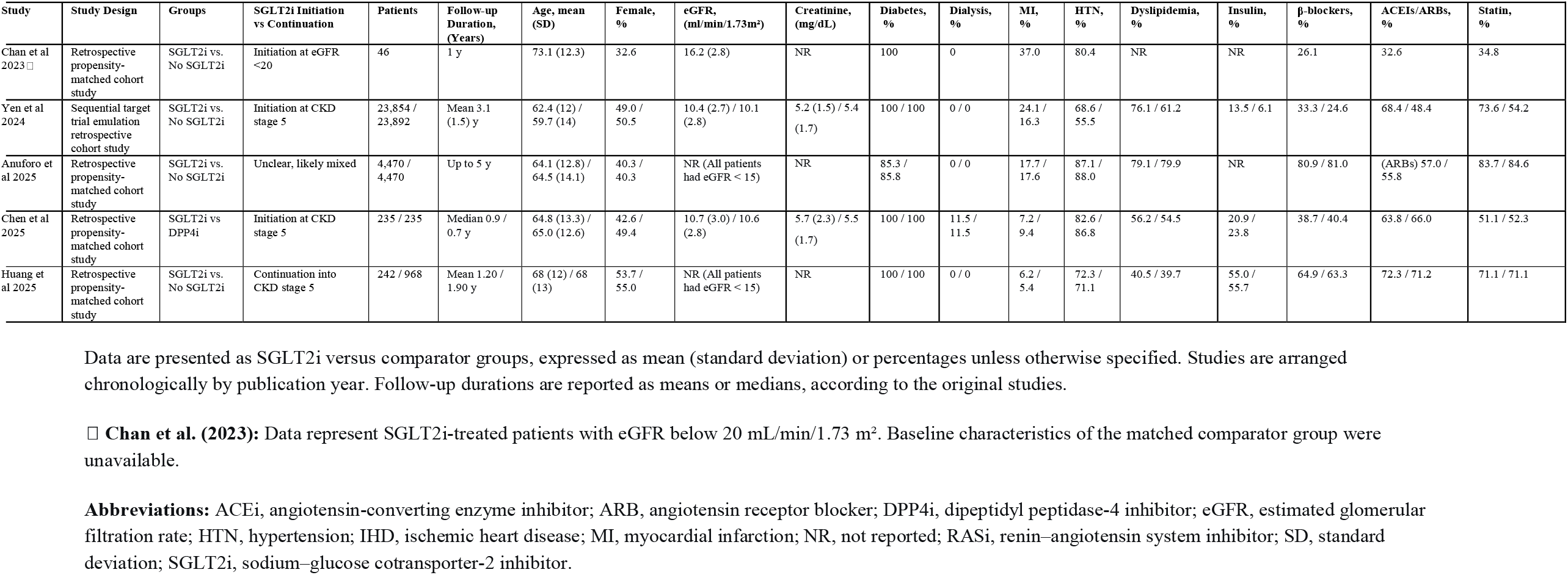
Baseline Characteristics of Studies Investigating SGLT2 Inhibitor Use in Patients with Stage 5 Chronic Kidney Disease.

| **Study** | **Study Design** | **Groups** | **SGLT2i Initiation vs Continuation** | **Patients** | **Follow-up Duration, (Years)** | **Age, mean (SD)** | **Female, %** | **eGFR, (ml/min/1.73m²)** | **Creatinine, (mg/dL)** | **Diabetes, %** | **Dialysis, %** | **MI, %** | **HTN, %** | **Dyslipidemia, %** | **Insulin, %** | **β-blockers, %** | **ACEIs/ARBs, %** | **Statin, %** |
|---|---|---|---|---|---|---|---|---|---|---|---|---|---|---|---|---|---|---|
| Chan et al 2023□ | Retrospective propensity-matched cohort study | SGLT2i vs. No SGLT2i | Initiation at eGFR <20 | 46 | 1 y | 73.1 (12.3) | 32.6 | 16.2 (2.8) | NR | 100 | 0 | 37.0 | 80.4 | NR | NR | 26.1 | 32.6 | 34.8 |
| Yen et al 2024 | Sequential target trial emulation retrospective cohort study | SGLT2i vs. No SGLT2i | Initiation at CKD stage 5 | 23,854 / 23,892 | Mean 3.1 (1.5) y | 62.4 (12) / 59.7 (14) | 49.0 / 50.5 | 10.4 (2.7) / 10.1 (2.8) | 5.2 (1.5) / 5.4 (1.7) | 100 / 100 | 0 / 0 | 24.1 / 16.3 | 68.6 / 55.5 | 76.1 / 61.2 | 13.5 / 6.1 | 33.3 / 24.6 | 68.4 / 48.4 | 73.6 / 54.2 |
| Anuforo et al 2025 | Retrospective propensity-matched cohort study | SGLT2i vs. No SGLT2i | Unclear, likely mixed | 4,470 / 4,470 | Up to 5 y | 64.1 (12.8) / 64.5 (14.1) | 40.3 / 40.3 | NR (All patients had eGFR < 15) | NR | 85.3 / 85.8 | 0 / 0 | 17.7 / 17.6 | 87.1 / 88.0 | 79.1 / 79.9 | NR | 80.9 / 81.0 | (ARBs) 57.0 / 55.8 | 83.7 / 84.6 |
| Chen et al 2025 | Retrospective propensity-matched cohort study | SGLT2i vs DPP4i | Initiation at CKD stage 5 | 235 / 235 | Median 0.9 / 0.7 y | 64.8 (13.3) / 65.0 (12.6) | 42.6 / 49.4 | 10.7 (3.0) / 10.6 (2.8) | 5.7 (2.3) / 5.5 (1.7) | 100 / 100 | 11.5 / 11.5 | 7.2 / 9.4 | 82.6 / 86.8 | 56.2 / 54.5 | 20.9 / 23.8 | 38.7 / 40.4 | 63.8 / 66.0 | 51.1 / 52.3 |
| Huang et al 2025 | Retrospective propensity-matched cohort study | SGLT2i vs. No SGLT2i | Continuation into CKD stage 5 | 242 / 968 | Mean 1.20 / 1.90 y | 68 (12) / 68 (13) | 53.7 / 55.0 | NR (All patients had eGFR < 15) | NR | 100 / 100 | 0 / 0 | 6.2 / 5.4 | 72.3 / 71.1 | 40.5 / 39.7 | 55.0 / 55.7 | 64.9 / 63.3 | 72.3 / 71.2 | 71.1 / 71.1 |

Data are presented as SGLT2i versus comparator groups, expressed as mean (standard deviation) or percentages unless otherwise specified. Studies are arranged chronologically by publication year. Follow-up durations are reported as means or medians, according to the original studies.
□ **Chan et al. (2023):** Data represent SGLT2i-treated patients with eGFR below 20 mL/min/1.73 m². Baseline characteristics of the matched comparator group were unavailable.
**Abbreviations:** ACEi, angiotensin-converting enzyme inhibitor; ARB, angiotensin receptor blocker; DPP4i, dipeptidyl peptidase-4 inhibitor; eGFR, estimated glomerular filtration rate; HTN, hypertension; IHD, ischemic heart disease; MI, myocardial infarction; NR, not reported; RASi, renin–angiotensin system inhibitor; SD, standard deviation; SGLT2i, sodium–glucose cotransporter-2 inhibitor.

Treatment exposure differed across studies. Yen (2024) and Chen (2025) examined SGLT2 inhibitor initiation in stage 5 CKD, while Chan (2023) evaluated initiation at eGFR <20 mL/min/1.73 m^2^ and required at least three months of treatment. Huang (2025) primarily examined treatment continuation into stage 5 CKD, whereas Anuforo (2025) did not specify when treatment began. Yen (2024) and Anuforo (2025) contributed the most statistical weight. ROBINS-I assessments identified serious risk of bias in three studies, moderate risk in one, and critical risk in one **(Figure S2)**.

SGLT2 inhibitor use was associated with fewer heart failure hospitalizations (HR, 0.78; 95% CI, 0.72–0.85; I^2^ = 0%) and acute myocardial infarctions (HR, 0.64; 95% CI, 0.56–0.72; I^2^ = 47%). Both associations remained significant after Hartung–Knapp adjustment. In contrast, associations with all-cause mortality (HR, 0.84; 95% CI, 0.58–1.21; I^2^ = 96%) and dialysis initiation (HR, 0.63; 95% CI, 0.30–1.33; I^2^ = 94%) were not statistically significant, with substantial heterogeneity for both outcomes.

When analyses were restricted to the two treatment-initiation cohorts, the association with fewer heart failure hospitalizations persisted (HR, 0.80; 95% CI, 0.74–0.87). However, associations with mortality (HR, 1.07; 95% CI, 0.87–1.31), dialysis initiation (HR, 0.66; 95% CI, 0.18–2.47), and myocardial infarction (HR, 0.88; 95% CI, 0.35–2.24) were not statistically significant.

Certainty of evidence was rated low for heart failure hospitalization and myocardial infarction and very low for mortality and dialysis initiation **(Table S2)**.

Two studies were included only in the qualitative synthesis. Huang (2025) reported fewer new-onset end-stage renal disease events (HR, 0.61), but no significant reduction in mortality or major adverse cardiac and cerebrovascular events (MACCE). Chan (2023) found fewer kidney-related events, defined as CKD progression, renal replacement therapy, or kidney-related death. However, the reduction in major adverse cardiovascular events (MACE) was not statistically significant after adjustment for multiple comparisons.

No study reported a statistically significant increase in adverse events among SGLT2 inhibitor users. Rates of ketoacidosis, urinary tract infections, hypoglycemia, acute kidney injury, and amputations were generally similar to or lower than those in comparator groups. Volume depletion was not reported.

## 4. DISCUSSION

In this meta-analysis of observational studies involving patients with stage 5 CKD, SGLT2 inhibitor use was associated with fewer heart failure hospitalizations and acute myocardial infarctions. Associations with mortality and dialysis initiation, however, remained uncertain. The lower rate of heart failure hospitalization is consistent with findings from our previous meta-analysis of randomized trials examining SGLT2 inhibitor initiation during hospitalization for acute heart failure ^10^. Because most of the pooled evidence came from treatment-initiation cohorts, these findings may be relevant to decisions about initiating SGLT2 inhibitors below current guideline thresholds.

Several limitations should be considered. Although the pooled analysis showed a significant association with fewer myocardial infarctions, this finding was no longer significant when the analysis was restricted to treatment-initiation cohorts, excluding the study with unclear exposure timing. All included studies were observational, leaving the possibility of residual confounding. In addition, the small number of studies, substantial heterogeneity for some outcomes, and concerns about risk of bias limit confidence in the estimates.

Overall, the available observational evidence suggests that SGLT2 inhibitors may offer cardiovascular benefits in stage 5 CKD, particularly by reducing heart failure hospitalizations. An association with fewer myocardial infarctions was also observed, although it was less robust in sensitivity analysis. Whether these agents improve survival or delay dialysis initiation remains unclear. Randomized trials, including the ongoing RENAL LIFECYCLE trial (NCT05374291), are needed to establish their benefits and risks and guide treatment initiation in patients with stage 5 CKD ^11^.

## Abbreviations

CI: confidence interval
CKD: chronic kidney disease
DPP-4: dipeptidyl peptidase-4
eGFR: estimated glomerular filtration rate
ESRD: end-stage renal disease
GRADE: Grading of Recommendations Assessment, Development and Evaluation
HR: hazard ratio
MI: myocardial infarction
MACE: major adverse cardiovascular events
MACCE: major adverse cardiac and cerebrovascular events
PRISMA: Preferred Reporting Items for Systematic Reviews and Meta-Analyses
PROSPERO: International Prospective Register of Systematic Reviews
REML: restricted maximum likelihood
ROBINS-I: Risk of Bias in Non-randomized Studies of Interventions
SGLT2: sodium–glucose cotransporter-2
SGLT2i: sodium–glucose cotransporter-2 inhibitor.

## Acknowledgments

None.

## DECLARATIONS

### Declaration of competing interest

The authors declare no conflicts of interest.

### Funding

No external funding was received for this study.

### Data Availability Statement

All data used in this study were derived from published articles and their supplementary materials. The extracted study-level data used for the meta-analysis are provided in the article and supplementary material.

### Ethics approval and informed consent

Ethics approval and informed consent were not required because this systematic review and meta-analysis used aggregate data from previously published studies and involved no direct participant contact.

### Author Contributions

**Hafiz M. Ahmed:** Conceptualization, Methodology, Project administration, Supervision, Investigation, Data curation; Formal analysis, Software, Visualization, Writing - original draft, Writing- review & editing.

**Ubaid ur Rehman:** Investigation, Data curation, Visualization, Writing - original draft, Writing - review & editing.

**Muhammad Owais:** Investigation, Data curation, Validation, Writing - original draft, Writing - review & editing.

## FIGURE LEGENDS

**Figure 1.**
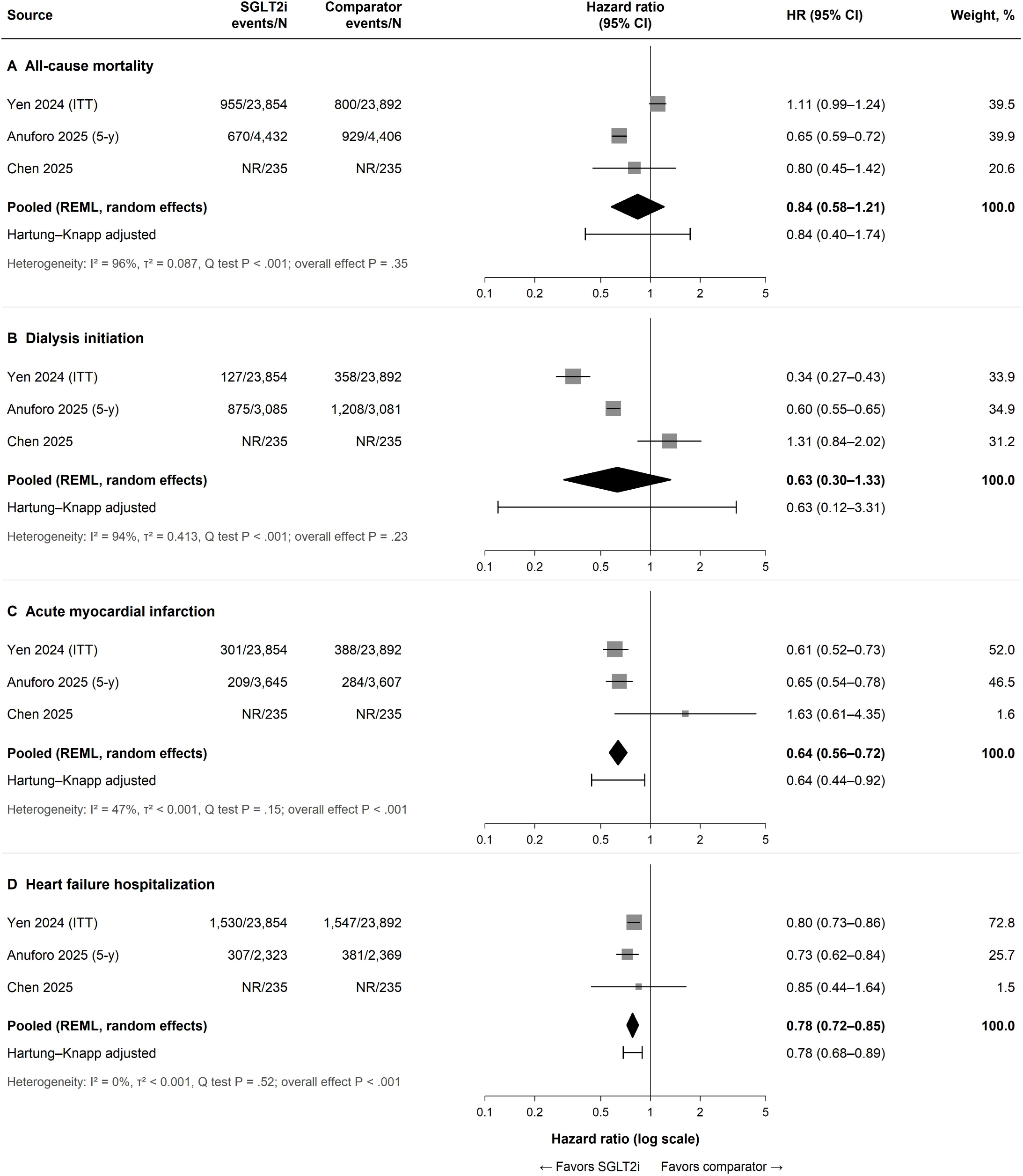
Association of SGLT2 Inhibitor Use With Clinical Outcomes in Stage 5 Chronic Kidney Disease. Forest plots show associations with (A) all-cause mortality, (B) dialysis initiation, (C) acute myocardial infarction, and (D) heart failure hospitalization. Hazard ratios (HRs) were pooled using inverse-variance random-effects models with restricted maximum likelihood (REML) estimation. Squares indicate study estimates (sized by weight), horizontal lines represent 95% confidence intervals (CIs), and diamonds indicate pooled estimates. Hartung–Knapp-adjusted CIs are shown below pooled estimates. Heterogeneity was assessed using I^2^. NR, not reported; SGLT2i, sodium–glucose cotransporter-2 inhibitor.

## Notes

### Competing Interest Statement

The authors have declared no competing interest.

### Author Declarations

This systematic review and meta-analysis exclusively used aggregate data from previously published, publicly available observational studies. All source data were available before the initiation of the review and were extracted from the original publications and their supplementary materials. No individual-level patient data or restricted-access databases were accessed. All analyses were conducted using published aggregate results.

