## Supplementary Material for "SGLT2 Inhibitors in Stage 5 Chronic Kidney Disease: A Systematic Review and Meta-analysis of Cardiovascular and Kidney Outcomes"

**Contents**

1. **Table S1.** Systematic Literature Search Strategy and Results by Database
2. **Figure S1.** PRISMA 2020 Flow Diagram of Study Identification, Screening, Eligibility, and Inclusion
3. **Figure S2.** Risk of Bias Assessment of Included Observational Studies Using the ROBINS-I Tool
4. **Table S2.** Absolute Effects and Certainty of Evidence for SGLT2 Inhibitor Therapy in Patients With Stage 5 Chronic Kidney Disease

**Table S1. Systematic Literature Search Strategy and Results by Database**

| **Database** | **Search Terms** | **Filters Applied** | **Results** |
| --- | --- | --- | --- |
| PubMed | ((("Chronic Kidney Disease") OR (CKD))) AND (((((("SGLT2 inhibitors")) OR (SGLT2i) OR (Empagliflozin)) OR (Dapagliflozin)) OR (Canagliflozin)) OR (Ertugliflozin)) OR (Sotagliflozin) | Age: Adult 19+ years | 939 |
| Scopus | TITLE-ABS-KEY ("Chronic Kidney Disease Stage 5" OR "CKD Stage 5") AND TITLE-ABS-KEY ("sodium glucose cotransporter 2" OR "sodium glucose transporter 2" OR SGLT2 OR SGLT-2 OR empagliflozin OR dapagliflozin OR canagliflozin OR ertugliflozin OR sotagliflozin) | Document Type: Articles | 383 |
| Cochrane Library | ("Chronic kidney disease stage 5" OR "ckd stage 5"):ti,ab,kw AND ("sodium glucose cotransporter 2" OR "SGLT-2" OR empagliflozin OR dapagliflozin OR canagliflozin OR ertugliflozin OR sotagliflozin):ti,ab,kw | Search in: Trials only | 170 |
| Google Scholar | "Chronic Kidney Disease" "SGLT2 Inhibitors" | First 200 results screened | 200 |

The systematic literature search was performed from database inception through April 10, 2026. Search strategies employed combinations of controlled vocabulary terms (MeSH in PubMed) and free-text keywords adapted to each database's syntax requirements. Boolean operators (AND, OR) were used to combine search concepts for chronic kidney disease and SGLT2 inhibitors. For Google Scholar, the first 200 results were screened due to relevance-ranking algorithms and diminishing returns beyond this threshold.

**Abbreviations:** CKD, chronic kidney disease; SGLT2, sodium-glucose cotransporter-2; SGLT2i, SGLT2 inhibitors; ti, title; ab, abstract; kw, keyword.

**Figure S1. PRISMA 2020 flow diagram of study identification, screening, eligibility, and inclusion**
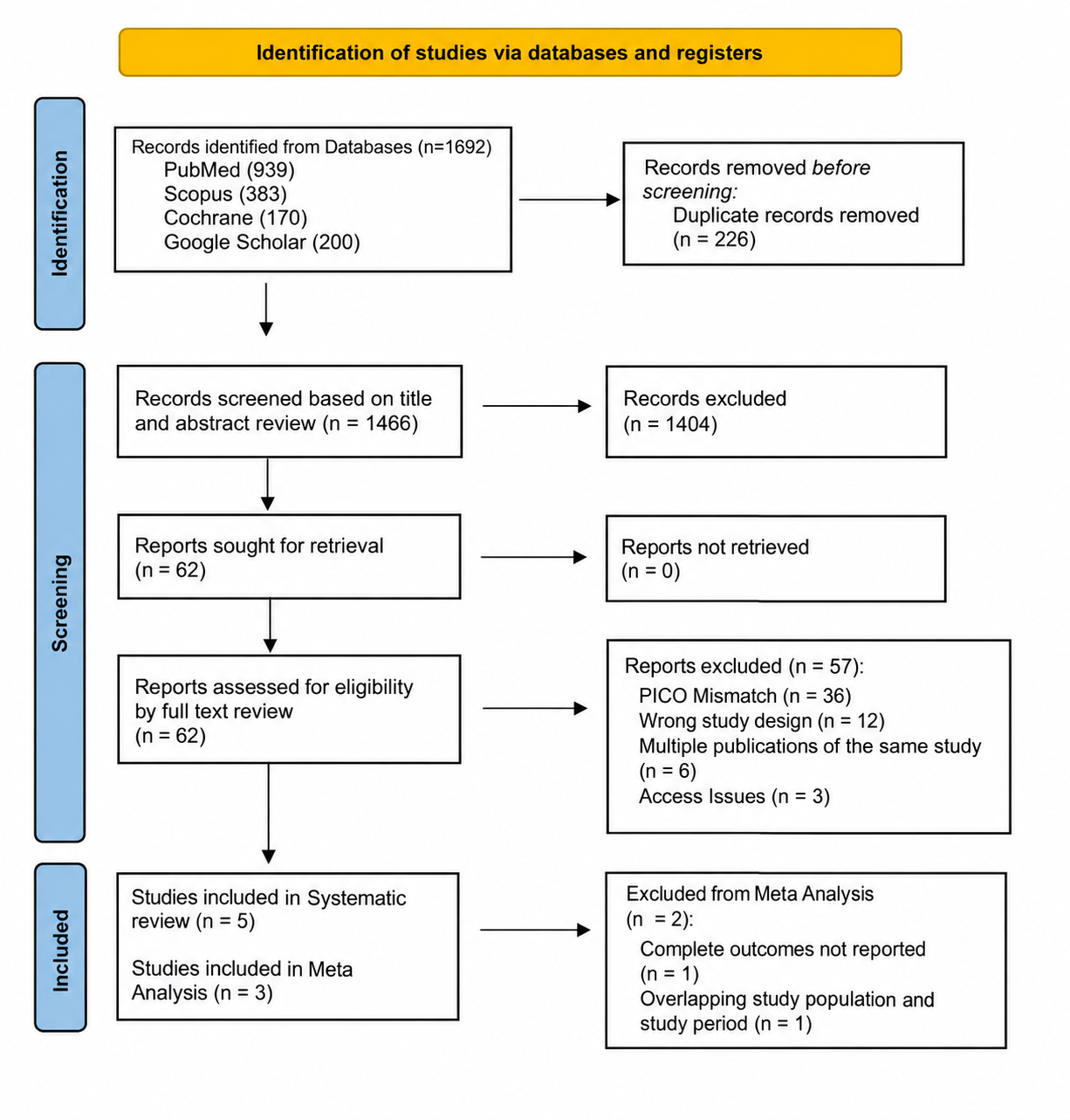
The flow diagram illustrates the study selection process according to PRISMA 2020 guidelines. Of 1,692 records identified, 226 duplicates were removed, and 1,466 records underwent title and abstract screening. Following full-text assessment of 62 reports, five studies were included in the systematic review, of which three were eligible for meta-analysis. Two studies were excluded from quantitative synthesis because of incomplete outcome reporting (n = 1) and overlapping study populations and periods (n = 1).

**Figure S2. Risk of bias assessment of included observational studies using the Risk of Bias in Non-randomized Studies of Interventions (ROBINS-I)**
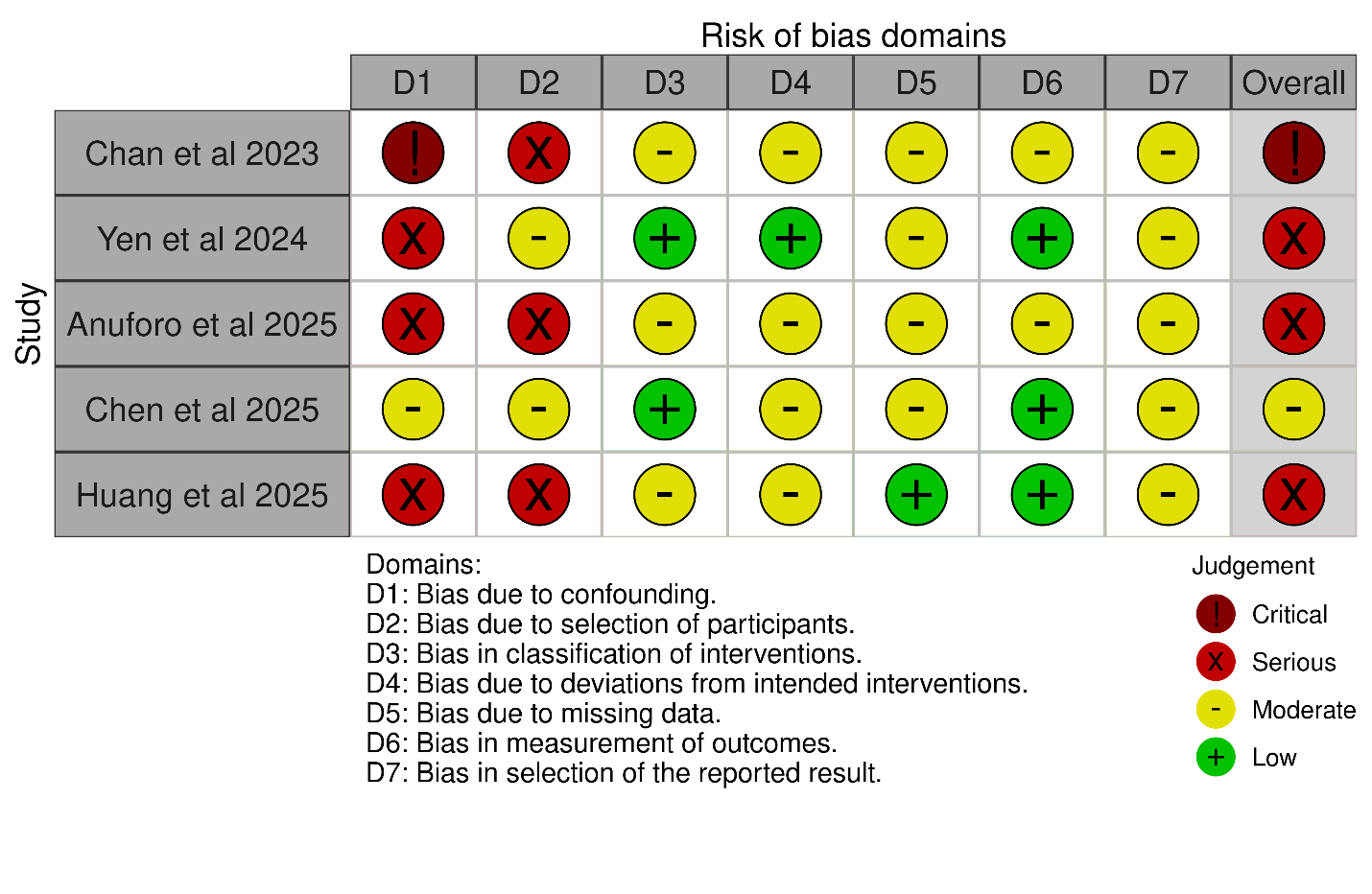

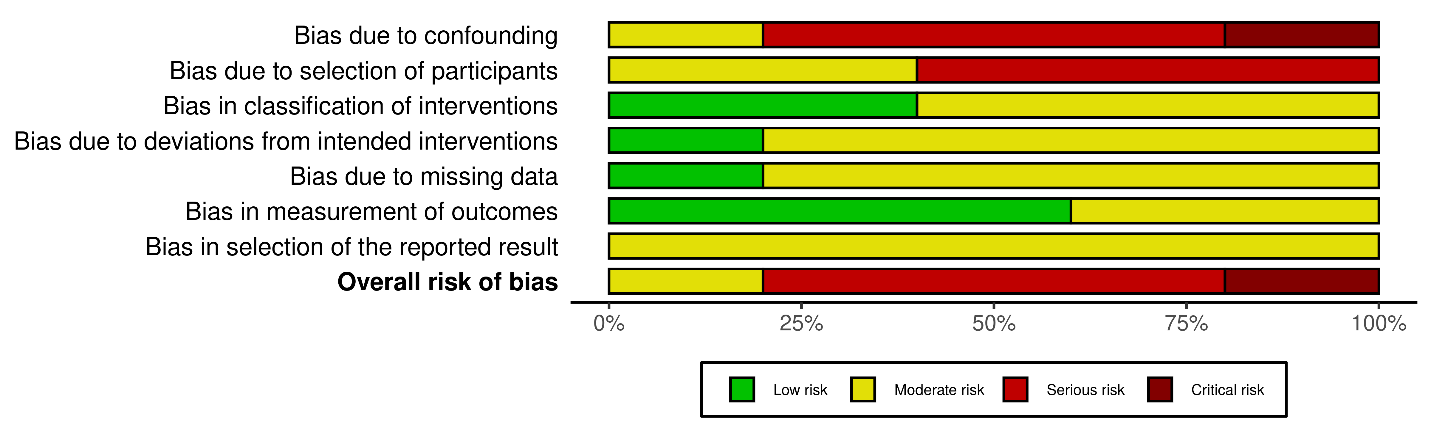
The upper panel shows study-level judgments across seven ROBINS-I domains, while the lower panel summarizes the proportion of studies in each risk category. Domains include confounding (D1), participant selection (D2), intervention classification (D3), deviations from intended interventions (D4), missing data (D5), outcome measurement (D6), and selective reporting (D7). Risk was classified as low (green), moderate (yellow), serious (red), or critical (dark red).

**Table S2. Absolute effects and certainty of evidence for SGLT2 inhibitor therapy in patients with stage 5 chronic kidney disease**

| **Outcome** | **Studies (patients), No.** | **HR (95% CI), REML random-effects model** | **HR (95% CI), Hartung–Knapp adjustment** | **I², %** | **SGLT2i risk, per 1000** | **Comparator risk, per 1000** | **Absolute difference per 1000 (95% CI)** | **NNT/NNH (95% CI)** | **Certainty (GRADE)** |
| --- | --- | --- | --- | --- | --- | --- | --- | --- | --- |
| Heart failure hospitalization | 3 (52,908) | 0.78 (0.72–0.85) | 0.78 (0.68–0.89) | 0 | 58 | 73 | 16 fewer (20 fewer to 11 fewer) | NNT 64 (50–91) | Lowᵃ |
| Acute myocardial infarction | 3 (55,468) | 0.64 (0.56–0.72) | 0.64 (0.44–0.92) | 47 | 16 | 24 | 9 fewer (11 fewer to 7 fewer) | NNT 114 (95–147) | Lowᵃ |
| All-cause mortality | 3 (57,054) | 0.84 (0.58–1.21) | 0.84 (0.40–1.74) | 96 | 52 | 61 | 10 fewer (25 fewer to 12 more) | Not estimable (CI includes no effect) | Very lowᵇ |
| Dialysis initiation | 3 (54,382) | 0.63 (0.30–1.33) | 0.63 (0.12–3.31) | 94 | 37 | 58 | 21 fewer (40 fewer to 18 more) | Not estimable (CI includes no effect) | Very lowᵇ |

Estimates were derived from three observational cohort studies. Hazard ratios were pooled using inverse-variance random-effects meta-analysis with restricted maximum likelihood (REML) estimation. Hartung–Knapp-adjusted confidence intervals (CIs) were reported in sensitivity analyses. Absolute risks, risk differences, and NNT/NNH were estimated rather than directly observed. Comparator risk was calculated from pooled event rates in the comparator groups of Yen (2024) and Anuforo (2025). Chen (2025) was excluded because only incidence rates were reported.

Certainty of Evidence was assessed using GRADE, starting at high certainty for observational studies evaluated with ROBINS-I and downgrading for risk of bias, inconsistency and imprecision, and indirectness.

**ᵃ** Downgraded two levels for risk of bias.

**ᵇ** Downgraded for risk of bias, inconsistency (I² ≥94%), and imprecision.

**Abbreviations:** CI, confidence interval; GRADE, Grading of Recommendations Assessment, Development and Evaluation; HR, hazard ratio; NNH, number needed to harm; NNT, number needed to treat; REML, restricted maximum likelihood; SGLT2i, sodium–glucose cotransporter-2 inhibitor.
